# Genetic Networks of Alzheimer’s Disease, Aging and Longevity in Humans

**DOI:** 10.1101/2023.01.06.23284252

**Authors:** Timothy Balmorez, Amy Sakazaki, Shin Murakami

## Abstract

Human genomic analysis and genome-wide association studies (GWAS) have identified genes that are risk factors for early and late-onset Alzheimer’s disease (AD genes). Although the genetics of aging and longevity has been extensively studied, previous studies have focused on a specific set of genes that have been shown to contribute to or to be a risk factor for AD. Thus, the connections among the genes involved in AD, aging, and longevity are not well understood. Here, we identified the genetic interaction networks (referred to as pathways) of aging and longevity within the context of AD, using a gene set enrichment analysis by Reactome that cross-references more than 100 bioinformatic databases to allow interpretation of the biological functions of gene sets through a wide variety of gene networks. We validated the pathways with a threshold of p-value < 1.00E-05, using the databases to extract lists of 356 AD genes, 307 aging-related (AR) genes, and 357 longevity genes. There was a broad range of biological pathways involved in AR and Longevity genes shared with AD genes. AR genes identified 261 pathways within the threshold of p < 1.00E-05, of which 26 pathways (10% of AR gene pathways) were further identified by overlapping genes among AD and AR genes. The overlapped pathways included Gene Expression (p = 4.05E-11) including ApoE, SOD2, TP53 and TGFB1 (p = 2.84E-10); Protein Metabolism and SUMOylation including E3 ligases and target proteins (p = 1.08E-07), ERBB4 signal transduction (p = 2.69E-06), Immune System including IL-3 and IL-13 (p = 3.83E-06), Programmed Cell Death (p = 4.36E-06) and platelet degranulation (p = 8.16E-06) among others. Longevity genes identified 49 pathways within the threshold, of which 12 pathways (24% of Longevity gene pathways) were further identified by overlapping genes among AD and Longevity genes. They include the immune system including IL-3 and IL-13 (p = 7.64E-08), plasma lipoprotein assembly, remodeling and clearance (p < 4.02E-06), metabolism of fat-soluble vitamins (p = 1.96E-05). Thus, this study provides shared genetic hallmarks of aging, longevity, and AD backed up by statistical significance. We discuss significant genes involved in these pathways, including TP53, FOXO, Sumoylation, IL4, IL6, APOE, and CEPT, and suggest that mapping the gene network pathways provide a useful basis for further medical research, education, and community outreach.

## Introduction

Alzheimer’s disease is the most frequent cause of dementia in which about 5.0 million people are living with AD in 2014 and the number is estimated to nearly triple by 2060 (Matthews et al., 2018). The centers for disease control and prevention (CDC) states that the number of people living with AD doubles every 5 years beyond age 65 (Accessed on January 2023; https://www.cdc.gov/aging/aginginfo/alzheimers.htm). Aging and genetic variations are well-known risks for AD. The genetic basis of AD has been characterized in the early onset of AD (EOAD), including amyloid precursor protein (APP) (Goate et al., 1991), presenilin 1 (PSEN1) (Sherrington et al., 1995), and presenilin 2 (PSEN2) (Rogaev et al., 1995), which account for less than 1% of AD cases (Sherva and Kwall, 2022). The vast majority of genetic risk factors fall into late-onset AD (LOAD). Additionally, the predictive contribution of each gene is modest for LOAD (Harold et al., 2009). Thus, research on the profiles of AD risk factor genes (AD genes) is expected to be effective. AD genes identified lipoprotein metabolism as a major hallmark of AD, which is tightly linked to a major mortality risk, cardiovascular disease (Vahdati Nia et al., 2017; Murakami and Lacayo, 2022). A component of lipoprotein metabolism includes ApoE isoforms which are well-known as a risk factor for LOAD (Mooser et al., 2000) as well as associated with longevity (Reviewed in Sebastiani et al., 2019). However, although there are genetic variants associated with aging (i.e., pathophysiological changes with increasing age) and longevity (i.e., length of lifespan), genetic interconnections with AD are not yet fully understood (Christensen et al., 2006; Vahdati Nia et al., 2017; Murakami and Lacayo, 2022). We reason that studying the gene sets or profiles is an effective way to understand AD and its relationship with aging and longevity. In this study, we chose a type of bioinformatic analysis using Reactome to elucidate the underlying molecular pathways and mechanisms.

Reactome is an open-source bioinformatic database with two functions. Firstly, it is a knowledge database of biological pathways manually curated and peer-reviewed, including twenty-seven biological pathway groups: they are autophagy, cell cycle, cell-cell communication, cellular responses to external stimuli, chromatin organization, circadian clock, developmental biology, digestion and absorption, disease, among others (Jassal et al., 2020). The pathway, reaction, and molecules pages extensively cross-reference to more than 100 bioinformatics resources, including ChEBI small molecule databases, Ensembl and UniProt databases, NCBI Gene, the UCSC Genome Browser, and PubMed. Secondly, it serves as a bioinformatic tool to perform gene set enrichment analysis (GSEA) (Milacic et al., 2012; Fabregat et al., 2018). GSEA is a computational method to identify and interpret biological functions of a gene set from genome-wide profiles. It uses gene annotations (such as gene ontology (GO), disease ontology (DO), and pathway annotations) and provides a ranked list of annotations with statistical significance for validation of the ranked list (Subramanian et al. 2005).

Using the Reactome analysis, we investigated three types of human genes: AD genes, aging-related (AR) genes, and Longevity genes. AD genes have been reported previously, which are validated based on meta-analyses of human GWAS (genome-wide association) studies (Bertram et al., 2007; Vahdati Nia et al., 2017; Murakami and Lacayo, 2022). Aging-related (AR) genes in humans are a list of genes identified based on an extensive literature review, followed by manually-curated annotation (de Magalhaes et al., 2009). The genes are from the meta-analysis study of human gene-expression analysis as well as predicted based on the studies in model systems, including the yeast, the nematode, the fruit fly, and the mouse (de Magalhaes et al., 2009). The proteomic and genomic map of the AR gene list has been reported earlier (de Magalhães and Toussaint, 2004). Longevity genes are the genes identified as gene variants associated with longevity (Budovsky et al., 2013). Notable risk factors for Alzheimer’s include APOE, as well as genes associated with inflammation and the insulin/IGF-1 signaling pathway (Willcox et al., 2008). However, although AD, aging, and longevity may be associated with each other, there is a lack of genetic interaction maps that overview each of them. Thus. we investigated the connections among the genes involved in AD, aging, and longevity.

## Materials and Methods

### Datasets

We used three sets of genes for our study: human Alzheimer’s Disease (AD) genes, Aging-Related (AR) genes, and Longevity genes. With these three gene sets, two additional gene sets were created by: (1) identifying the genes shared both by the AD genes and by the AR genes (AD-AR Overlap), and (2) identifying the genes shared both by AD genes and Longevity genes (AD-Longevity Overlap). Firstly, we utilized the AlzGene database (www.alzgene.org; last accessed May 2021) to extract a list of 680 identified human AD genes from GWAS and previous linkage studies (Bertram et al., 2007; Olgiati et al., 2011). Each gene from the database was linked to a number of positive or negative test results from each GWAS study, which were then used to validate the reliability of the data. Out of the 680 genes, 356 genes had positive results (AD genes) and 324 genes had negative results. Since the definite negative and positive results were not included in the database, we used the *p*-value from previous studies. If *p* < 0.05, we assumed the data was reliable and counted them as a positive result; studies with *p* > 0.05 were regarded as negative and were not included in our study. Research outcomes listed as trends or inconclusive were also not included in our study. Secondly, we utilized the GenAge database (www.genomics.senescence.info; last accessed May 2021), a benchmark database of 307 genes possibly related to human aging (AR). The genes were extensively reviewed for inclusion based on findings in model organisms put in the context of human biology plus genes directly related to aging in humans (Tacutu et al., 2018). Thirdly, we utilized the LongevityMap database (www.genomics.senescence.info/longevity/; last accessed December 2021), a database of 751 genes associated with human longevity (Budovsky et el., 2013). In the database, 394 genes were labeled “non-significant” and thus excluded from this study for analysis. The remaining 357 genes were labeled as “significant” (Longevity genes) and used in this study. Finally, the list of AD Genes was used to cross-reference the presence of any genes shared by the combinations of each gene sets (AD-AR overlap genes and AD-Longevity overlap genes). When comparing AD genes with AR genes, 41 overlappings (AD-AR overlap) genes were found. When comparing AD genes and Longevity genes, 43 overlappings (AD-Longevity overlap) genes were found.

### Gene Ontology: Reactome Analysis

We used Reactome (www.reactome.org) to analyze the pathways involved in each gene set. The pathways with a threshold of p < 10E-5 were selected and used for this study. Reactome analysis was performed as described previously (Vahdati Nia et al., 2017; Murakami and Lacayo, 2022). Reactome FIViz was used to determine enrichment in the Functional Interaction (FI) network, the pathway enrichment of the genes of interest, followed by converting the results to interactomes. Statistics and false discovery rate (FDR) were calculated by the Reactome FIViz. We used Cytoscape ver. 3.8.2 (Java version: 11.0.6) to run the Reactome software plugin, Reactome FIViz app (Wu et al., 2014). The version of the pathway database was Reactome v76 (released on March 21, 2021; last accessed on May 26, 2021).

## Results

Figure 1 summarises the overall analysis of the gene sets. We identified five gene sets as described in the method. They are (1) 356 positive AD genes; (2) 307 AR genes; (3) 357 Longevity genes; (4) 41 AD-AR overlap genes; and (5) 43 AD-Longevity overlap genes (Figure 1). We then perform a Reactome analysis of each gene set that generated biological pathways relevant to the gene set. The pathways were validated by using the threshold of p < 10E-5; in other words, we excluded the pathways with a false discovery rate of more than 10E-3. In each Table, we included the top ten results from each Reactome analysis to be included in our results. We further identified specific Reactome groups from within the generalized groups.

**Figure 1.**
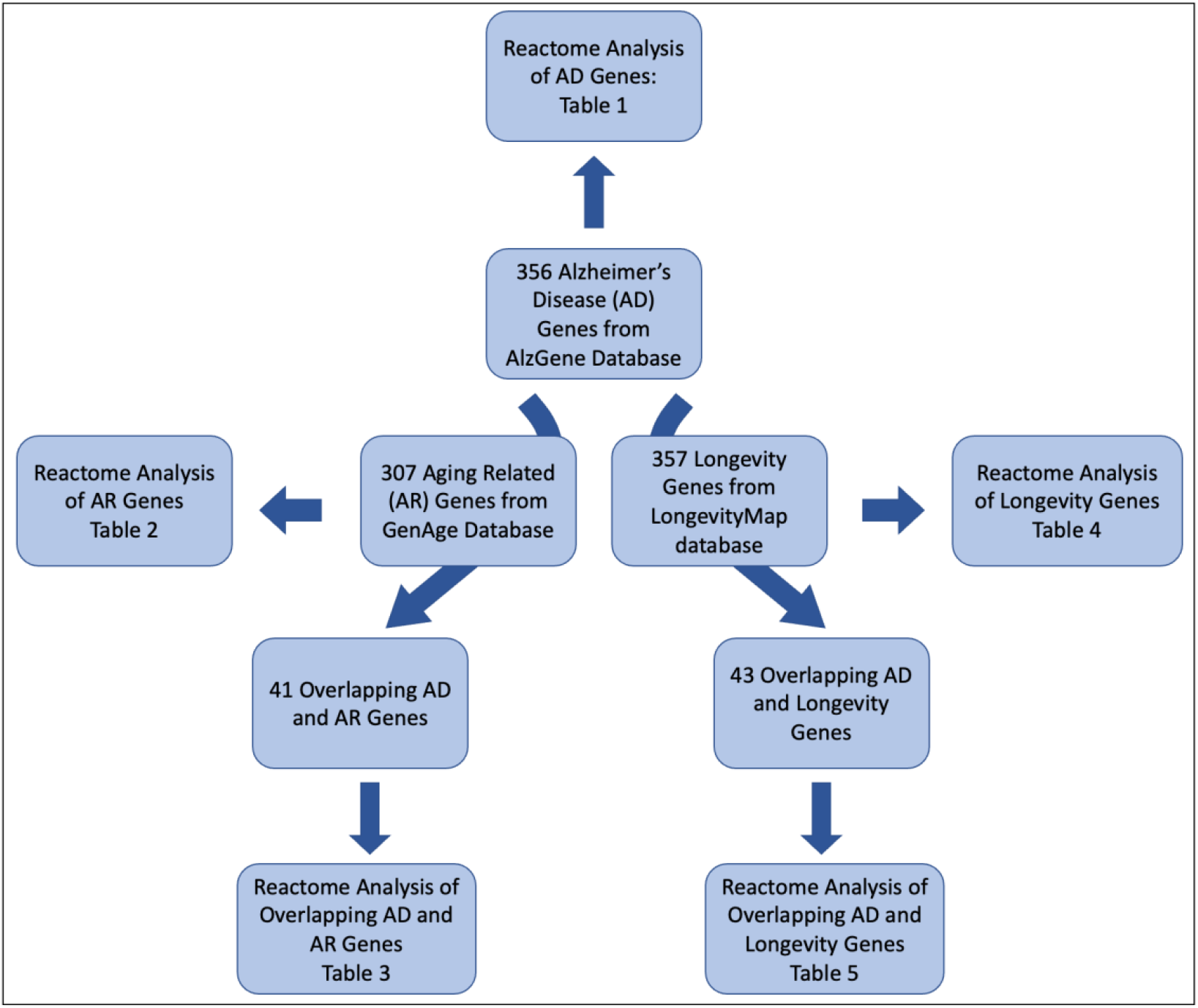
Schematic providing a breakdown of the methodology used in this study. We used Reactome analysis for AD genes, AR genes, Longevity genes, overlapping AD and AR genes, and overlapping AD and Longevity genes.

### AD Genes

The Reactome analysis of the AD genes identified 161 pathways. Of them, 53 pathways showed p < 1.0E-05, which is roughly equivalent to the false discovery rate < 1.0E-03. The significant pathways can be summarized into the following three general Reactome pathway groups: Metabolism of RNA, Transport of Small molecules, and Immune System. The top 10 results are shown in Table 1.

**TABLE 1.** Reactome pathway enrichment analysis results - Positive AlzGene (AD) genes.

| Reactome Pathway | No. of total proteins in pathway | No. of hits in pathway | p-value | FDR value | Hit gene |
| --- | --- | --- | --- | --- | --- |
| tRNA processing in the mitochondrion | 42 | 19 | 1.11E-16 | 1.01E-13 | *MT-ND6, MT-TQ, MT-ND4L, MT-ND4, MT-CO1, MT-TT, MT-TR, MT-ND2, MT-ND3, MT-ND1, MT-TH, MT-CO2, MT-TG, MT-CO3, MT-TS2, MT-ATP6, MT-ATP8, MT-RNR1, MT-CYB* |
| Plasma lipoprotein assembly, remodeling, and clearance | 71 | 21 | 4.44E-16 | 1.34E-13 | *LIPA, LIPC, SOAT1, CETP, APOE, A2M, ABCA1, VLDLR, LDLR, NR1H2, ABCG1, LPL, ALB, APOA1, APOA4, APOA5, NPC1, NPC2, APOC4, APOC2, APOC1* |
| rRNA processing in the mitochondrion | 38 | 17 | 4.44E-16 | 1.34E-13 | *MT-ND4L, MT-ND4, MT-CO1, MT-TT, MT-TR, MT-ND2, MT-ND3, MT-ND1, MT-TH, MT-CO2, MT-TG, MT-CO3, MT-TS2, MT-ATP6, MT-ATP8, MT-RNR1, MT-CYB* |
| Interleukin-4 and Interleukin-13 signaling | 112 | 21 | 2.57E-12 | 5.80E-10 | *ICAM1, TP53, MAOA, PIK3R1, HMOX1, CD36, IL10, IL18, IL1A, IL1B, PTGS2, ALOX5, F13A1, TNF, TGFB1, POU2F1, IL6, IL8, MMP1, MMP3, CCL2* |
| Plasma lipoprotein clearance | 33 | 12 | 9.58E-11 | 1.73E-08 | *LIPA, LIPC, SOAT1, APOE, VLDLR, LDLR, NR1H2, APOA1, NPC1, NPC2, APOC4, APOC1* |
| Interleukin-10 signaling | 47 | 13 | 4.28E-10 | 6.46E-08 | *IL1RN, ICAM1, CCR2, IL10, IL18, IL1A, IL1B, PTGS2, TNF, IL6, IL8, CCL3, CCL2* |
| Retinoid metabolism and transport | 44 | 12 | 2.36E-09 | 3.05E-07 | *LRAT, HSPG2, APOE, LDLR, LPL, APOA1, APOA4, LRP1, LRP2, LRP8, TTR, APOC2* |
| Metabolism of fat-soluble vitamins | 48 | 12 | 6.14E-09 | 6.93E-07 | *LRAT, HSPG2, APOE, LDLR, LPL, APOA1, APOA4, LRP1, LRP2, LRP8, TTR, APOC2* |
| tRNA processing | 146 | 19 | 1.09E-08 | 1.09E-06 | *MT-ND6, MT-TQ, MT-ND4L, MT-ND4, MT-CO1, MT-TT, MT-TR, MT-ND2, MT-ND3, MT-ND1, MT-TH, MT-CO2, MT-TG, MT-CO3, MT-TS2, MT-ATP6, MT-ATP8, MT-RNR1, MT-CYB* |
| Plasma lipoprotein remodeling | 32 | 10 | 1.47E-08 | 1.32E-06 | *LIPC, CETP, APOE, ABCG1, LPL, ALB, APOA1, APOA4, APOA5, APOC2* |

The top hit for the enriched pathway analysis (of AD positive genes) was “tRNA processing in the mitochondrion” (FDR = 1.01E-13; p = 1.11E-16), which was a sub-pathway topic under “tRNA processing” (FDR = 1.09E-6; p = 1.09E-8), which was a sub-pathway topic under “metabolism of RNA” in the hierarchy panel. “tRNA processing in the mitochondrion” involves 42 proteins and 19 of those proteins were found to be shared in our positively tested AD genes (FDR = 1.01E-13; p = 1.11E-16). The second top hit was “Plasma lipoprotein assembly, remodeling, and clearance” (FDR = 1.34E-1; p= 4.44E-16) from the “Transport of Small Molecules” general pathway. The third-fourth top hit, “Interleukin-4 and Interleukin-13 signaling” (FDR = 5.80E-10; p= 2.57E-12) is a part of the “Immune System” general pathway.

### AR Genes

669 pathways were identified by the Reactome analysis of the 307 AR genes. Using the threshold, we validated the pathways and selected 261 pathways using the threshold of *p* < 1.0E-05. The significant pathways identified by the AR genes fell into the following six general Reactome pathway groups: Immune System, Signal Transduction, Metabolism of Proteins, Gene Expression, Cellular Responses to External Stimuli, and DNA repair. The top ten results are shown in Table 2.

**TABLE 2.** Reactome pathway enrichment analysis results - GenAge (AR) genes.

| **Reactome Pathway** | **No. of total proteins in pathway** | **No. of hits in pathway** | **p-value** | **FDR value** | **Hit gene** |
| --- | --- | --- | --- | --- | --- |
| Signaling by Interleukins | 452 | 59 | 1.11E-16 | 8.66E-15 | *APP, ATF2, MYC, AKT1, MIF, TP53, PIK3R1, HIF1A, STAT5A, STAT5B, JUN, GRB2, CDKN1A, IKBKB, JAK2, FOS, CTF1, PTGS2, RELA, STAT3, VEGFA, IRS1, IRS2, TNF, SQSTM1, SHC1, FOXO3, FOXO1, SOCS2, UBB, HSPA9, HSPA8, TGFB1, IL2, NFKB1, NFKB2, NFKBIA, IL6, IL7, BCL2, IL7R, HMGB1, PIK3CB, LMNB1, HSP90AA1, YWHAZ, CREB1, PIK3CA, IL2RG, CDC42, MAPK9, MAPK8, PTK2B, MAPK3, PTPN11, MAPK14, S100B, SOD2, SOD1* |
| DNA Double-Strand Break Repair | 149 | 30 | 1.11E-16 | 8.66E-15 | *CHEK2, TP53, PRKDC, TP53BP1, XRCC6, XRCC5, ATM, ATR, FEN1, BRCA1, BRCA2, PARP1, SIRT6, PCNA, WRN, RPA1, NBN, BLM, UBB, ABL1, UBE2I, SUMO1, H2AFX, CCNA2, MAPK8, POLD1, RAD52, RAD51, ERCC4, ERCC1* |
| PIP3 activates AKT signaling | 286 | 46 | 1.11E-16 | 8.66E-15 | *ATF2, AKT1, TP53, PDGFB, PIK3R1, EGR1, JUN, INSR, GRB2, CDKN1A, BMI1, PPARG, EGFR, RICTOR, IRS1, IRS2, PDGFRB, PDGFRA, FOXO4, FOXO3, FOXO1, UBB, FGF23, KL, ESR1, MDM2, FGFR1, GSK3B, GSK3A, PTEN, PIK3CB, PDPK1, NRG1, CREB1, PIK3CA, HDAC2, HDAC3, HDAC1, INS, ERBB2, MAPK3, EGF, PTPN11, MTOR, PML, STUB1* |
| Intracellular signaling by second messengers | 326 | 49 | 1.11E-16 | 8.66E-15 | *ATF2, AKT1, PRKCD, PRKCA, TP53, PDGFB, PIK3R1, EGR1, JUN, INSR, GRB2, CDKN1A, BMI1, PPARG, EGFR, ADCY5, RICTOR, IRS1, IRS2, PDGFRB, PDGFRA, FOXO4, FOXO3, FOXO1, UBB, FGF23, KL, ESR1, MDM2, FGFR1, GSK3B, GSK3A, PTEN, PIK3CB, PDPK1, NRG1, CREB1, PIK3CA, HDAC2, HDAC3, HDAC1, INS, ERBB2, MAPK3, EGF, PTPN11, MTOR, PML, STUB1* |
| Signaling by Receptor Tyrosine Kinases | 493 | 60 | 1.11E-16 | 8.66E-15 | *ATF2, AKT1, PRKCD, PRKCA, PDGFB, PIK3R1, APOE, STAT5A, EGR1, STAT5B, JUND, INSR, IGF2, IGF1, PTK2, GRB2, JAK2, HRAS, FOS, NCOR1, EGFR, RICTOR, STAT3, VEGFA, CTNNB1, IRS1, IRS2, IGF1R, EP300, PDGFRB, PDGFRA, SHC1, PSEN1, UBB, FGF23, KL, BDNF, ESR1, FGFR1, FLT1, PIK3CB, HSP90AA1, PDPK1, NRG1, NGF, CREB1, PIK3CA, INS, CDC42, ERBB2, PTK2B, MAPK3, PTPN1, EGF, PTPN11, MAPK14, S100B, MTOR, BAX, STUB1* |
| FOXO-mediated transcription | 66 | 22 | 1.11E-16 | 8.66E-15 | *AKT1, DDIT3, TXN, PCK1, PPARGC1A, CREBBP, CDKN1A, SIRT1, SIRT3, NR3C1, STK11, EP300, FOXO4, FOXO3, FOXO1, SIN3A, YWHAZ, CAT, HDAC2, HDAC1, INS, SOD2* |
| RNA Polymerase II Transcription | 1363 | 110 | 1.11E-16 | 8.66E-15 | *RB1, ATF2, MT-CO1, SERPINE1, CHEK2, MYC, AKT1, TP63, MED1, AR, DDIT3, TP53, TXN, PRDX1, APOE, PCK1, PPARGC1A, CREBBP, JUN, ATM, ATR, TP73, CDKN1A, BRCA1, BMI1, PARP1, FOS, SIRT1, SIRT3, NCOR2, NCOR1, PPARG, PPARA, PCNA, HTT, EGFR, RELA, WRN, RICTOR, CDKN2B, CDKN2A, GSR, RPA1, GTF2H2, MLH1, VEGFA, CTNNB1, FAS, NR3C1, CTGF, STK11, EP300, NBN, IGFBP3, BLM, MAX, FOXO4, FOXO3, FOXO1, UBB, ABL1, TFAP2A, TGFB1, UBE2I, BDNF, ESR1, IL2, NFKB1, CDK7, IL6, SP1, CDK1, MDM2, PIN1, TCF3, GSK3B, PTEN, SUMO1, SIN3A, TBP, PDPK1, H2AFX, YWHAZ, CCNA2, CREB1, TFDP1, CAT, CEBPB, HDAC2, HDAC3, HDAC1, PPM1D, HSPD1, INS, ERBB2, E2F1, MAPK3, PTPN1, PTPN11, MAPK14, SOD2, MTOR, PML, RAD51, ERCC3, SST, ERCC2, BAX, STUB1, TAF1* |
| Generic Transcription Pathway | 1234 | 110 | 1.11E-16 | 8.66E-15 | *RB1, ATF2, MT-CO1, SERPINE1, CHEK2, MYC, AKT1, TP63, MED1, AR, DDIT3, TP53, TXN, PRDX1, APOE, PCK1, PPARGC1A, CREBBP, JUN, ATM, ATR, TP73, CDKN1A, BRCA1, BMI1, PARP1, FOS, SIRT1, SIRT3, NCOR2, NCOR1, PPARG, PPARA, PCNA, HTT, EGFR, RELA, WRN, RICTOR, CDKN2B, CDKN2A, GSR, RPA1, GTF2H2, MLH1, VEGFA, CTNNB1, FAS, NR3C1, CTGF, STK11, EP300, NBN, IGFBP3, BLM, MAX, FOXO4, FOXO3, FOXO1, UBB, ABL1, TFAP2A, TGFB1, UBE2I, BDNF, ESR1, IL2, NFKB1, CDK7, IL6, SP1, CDK1, MDM2, PIN1, TCF3, GSK3B, PTEN, SUMO1, SIN3A, TBP, PDPK1, H2AFX, YWHAZ, CCNA2, CREB1, TFDP1, CAT, CEBPB, HDAC2, HDAC3, HDAC1, PPM1D, HSPD1, INS, ERBB2, E2F1, MAPK3, PTPN1, PTPN11, MAPK14, SOD2, MTOR, PML, RAD51, ERCC3, SST, ERCC2, BAX, STUB1, TAF1* |
| Cellular responses to stress | 554 | 61 | 1.11E-16 | 8.66E-15 | *RB1, ATF2, AR, DDIT3, TP53, TXN, HIF1A, PRDX1, CREBBP, JUN, ATM, ATR, CDKN1A, BMI1, FOS, TERF1, SIRT1, TERF2, GSTP1, RELA, HSF1, RAE1, CDKN2B, CDKN2A, IFNB1, GSR, STAT3, RPA1, VEGFA, HSPA1B, HSPA1A, NR3C1, EP300, NBN, MAP3K5, EEF1A1, VCP, UBB, HSPA9, HSPA8, NFKB1, IL6, SP1, MDM2, GSK3B, LMNB1, HSP90AA1, GPX1, H2AFX, CCNA2, TFDP1, CAT, CEBPB, MAPK9, MAPK8, E2F1, MAPK3, MAPK14, SOD2, MTOR, SOD1* |
| SUMO E3 ligases<br>SUMOylate target proteins | 169 | 36 | 1.11E-16 | 8.66E-15 | *AR, TP53, TP53BP1, PPARGC1A, CREBBP, TOP2A, TOP2B, BRCA1, BMI1, PARP1, NCOR2, PPARG, PPARA, PCNA, RELA, WRN, RAE1, CDKN2A, RPA1, NR3C1, EP300, HIC1, BLM, TFAP2A, UBE2I, ESR1, NFKB2, NFKBIA, MDM2, SUMO1, SIN3A, TOP1, HDAC2, HDAC1, PML, RAD52* |

The top hits for the enriched pathway analysis and their general pathways were the following: Signaling by interleukins (Immune system), DNA Double-Strand Break Repair (DNA repair), PIP3 activates AKT Signaling (Signal Transduction), FOXO-mediated transcription (Gene Expression), cellular responses to stress (cellular responses to external stimuli), SUMO E3 ligases SUMOylate target proteins (Metabolism of Proteins), cellular senescence (cellular responses to external stimuli), transcriptional regulation by TP53 (Gene Expression), and SUMOylation (Protein Metabolism). All results had an FDR = 8.66E-15; *p* = 1.11E-16.

### Longevity Genes

34 pathways were identified in the Reactome analysis of 357 Longevity genes. Of them, we validated and selected 12 pathways that satisfied the threshold of p < 1.0E-05. The significant pathways resulted in two general pathways: Signal Transduction and Gene Expression. The top 10 results are shown in Table 3. The top results in the enrichment pathway analysis were part of the “Signal Transduction” general pathway. The Reactome pathway “MTOR signaling”, (FDR = 5.15E-14; p = 1.11E-16) is a sub-pathway topic under “intracellular signaling by second messengers” (FDR = 5.39E-12, p = 1.74E-14). “MTOR signaling pathway” involves 41 proteins and 19 of those proteins were found to be shared in our Longevity genes. “PIP3 activates AKT signaling”, (FDR = 2.39E-11, p = 1.03E-13) is also a sub-pathway topic under the same pathway. In the “Gene Expression” general pathway, the significant pathways were “RNA Polymerase II Transcription” (FDR = 1.29E-09, p = 1.04E-11) and two sub-pathways – “TP53 Regulates Metabolic Genes” (FDR = 1.29E-09, p = 1.11E-11) and “FOXO-mediated transcription” (FDR = 1.78E-09, p = 1.73E-11).

**TABLE 3.** Reactome pathway enrichment analysis results - Longevity genes.

| Reactome Pathway | No. of Total Proteins in Pathway | No. of hits in pathway | p-value | FDR | HitGenes |
| --- | --- | --- | --- | --- | --- |
| mTOR signalling | 41 | 19 | 1.11E-16 | 5.15E-14 | *AKT1, RRAGA, RRAGC, RRAGB, RRAGD, RPTOR, LAMTOR2, LAMTOR3, RPS6, TSC2, TSC1, EIF4EBP1, RHEB, MLST8, AKT1S1, EIF4E, EIF4B, MTOR, RPS6KB1* |
| mTORC1-mediated signalling | 24 | 16 | 1.11E-16 | 5.15E-14 | *RRAGA, RRAGC, RRAGB, RRAGD, RPTOR, LAMTOR2, LAMTOR3, RPS6, EIF4EBP1, RHEB, MLST8, AKT1S1, EIF4E, EIF4B, MTOR, RPS6KB1* |
| Intracellular signaling by second messengers | 326 | 34 | 1.74E-14 | 5.39E-12 | *AKT1, PRKCA, RRAGA, RRAGC, RRAGB, RRAGD, TP53, MAPKAP1, TGFA, INSR, CDKN1A, RPTOR, PPARG, EGFR, RICTOR, CAMK4, LAMTOR2, LAMTOR3, IRS2, TSC2, FOXO4, FOXO3, FOXO1, KL, ESR1, NBEA, RHEB, FGFR1, PRR5, MLST8,* |
|  |  |  |  |  | *PIK3CA, AKT1S1, INS, MTOR* |
| PIP3 activates AKT signaling | 286 | 31 | 1.03E-13 | 2.39E-11 | *AKT1, RRAGA, RRAGC, RRAGB, RRAGD, TP53, MAPKAP1, TGFA, INSR, CDKN1A, RPTOR, PPARG, EGFR, RICTOR, LAMTOR2, LAMTOR3, IRS2, TSC2, FOXO4, FOXO3, FOXO1, KL, ESR1, RHEB, FGFR1, PRR5, MLST8, PIK3CA, AKT1S1, INS, MTOR* |
| Generic Transcription Pathway | 1234 | 66 | 3.56E-13 | 6.59E-11 | *SERPINE1, AKT1, RUNX3, RRAGA, RRAGC, RRAGB, RRAGD, TP53, MAPKAP1, TGFA, GATA4, APOE, ATRIP, PPARGC1A, ATM, CDKN1A, MSTN, RPTOR, SREBF1, SIRT1, SIRT3, RAD51D, RARB, PPARG, SGK1, EGFR, WRN, RICTOR, WWOX, CDKN2B, GSR, MLH1, VEGFA, CAMK4, FAS, LAMTOR2, LAMTOR3, NR3C1, YY1, KCTD1, TSC2, TSC1, TBL1XR1, CSF1R, FOXO4, FOXO3, FOXO1, PLXNA4, TGFB1, ESRRG, ESR1, NFKB1, IL6, CDK6, RHEB, PRR5, EXO1, MLST8, YWHAG, H2AFX, IFNG, INS, TXNRD1, SOD2, MTOR, ERCC2* |
| Energy dependent regulation of mTOR by LKB1-AMPK | 29 | 12 | 2.20E-12 | 3.39E-10 | *RRAGA, RRAGC, RRAGB, RRAGD, RPTOR, LAMTOR2, LAMTOR3, TSC2, TSC1, RHEB, MLST8, MTOR* |
| RNA Polymerase II Transcription | 1363 | 67 | 1.04E-11 | 1.29E-09 | *SERPINE1, AKT1, RUNX3, RRAGA, RRAGC, RRAGB, RRAGD, TP53, MAPKAP1, TGFA, GATA4, APOE, ATRIP, PPARGC1A, ATM, CDKN1A, MSTN, RPTOR, SREBF1, SIRT1, SIRT3, RAD51D, RARB, PPARG, SGK1, EGFR, WRN, RICTOR, WWOX, CDKN2B, GSR, MLH1, VEGFA, CAMK4, FAS, LAMTOR2, LAMTOR3, NR3C1, YY1, KCTD1, TSC2, TSC1, TBL1XR1, CSF1R, FOXO4, FOXO3, FOXO1, PLXNA4, TGFB1, ESRRG, ESR1, NFKB1, IL6, CDK6, RHEB, PRR5, EXO1, MLST8, YWHAG, H2AFX, IFNG, POLDIP3, INS, TXNRD1, SOD2,* |
|  |  |  |  |  | *MTOR, ERCC2* |
| TP53 Regulates Metabolic Genes | 89 | 17 | 1.11E-11 | 1.29E-09 | *AKT1, RRAGA, RRAGC, RRAGB, RRAGD, TP53, RPTOR, GSR, LAMTOR2, LAMTOR3, TSC2, TSC1, RHEB, MLST8, YWHAG, TXNRD1, MTOR* |
| FOXO-mediated transcription | 66 | 15 | 1.73E-11 | 1.78E-09 | *AKT1, PPARGC1A, CDKN1A, MSTN, SREBF1, SIRT1, SIRT3, NR3C1, FOXO4, FOXO3, FOXO1, PLXNA4, YWHAG, INS, SOD2* |
| Transcriptional Regulation by TP53 | 368 | 30 | 2.68E-10 | 2.25E-08 | *AKT1,RRAGA,RRAGC,RRAGB,RRAGD,TP53,MAPK AP1,ATRIP,ATM,CDKN1A,RPTOR,RAD51D,SGK1, WRN,RICTOR,GSR,MLH1,FAS,LAMTOR2,LAMTOR 3,TSC2,TSC1,RHEB,PRR5,EXO1,MLST8,YWHAG,T XNRD1,MTOR,ERCC2* |

### AD-AR overlap genes

41 AR genes (13% of the genes) overlapped with AD genes (AD-AR overlap genes) (Table 4). The Reactome analysis of the AD-AR overlaps genes 261 pathways. Of them, we validated and selected 24 pathways that satisfied the threshold of p < 1.0E-05. The pathways resulted in five general Reactome groups: Gene Expression, Metabolism of Proteins, Programmed Cell Death, Signal Transduction, and Immune System. The top ten results are shown in Table 5.

**TABLE 4.** Comparison of AR and positive AD genes.

| **GenAge (AR) Genes** | **AlzGene (AD) Genes** | **Overlap Genes** | **Overlapped in AR genes (%)** |
| --- | --- | --- | --- |
| 307 | 356 | 41 | 13% |

**TABLE 5.**
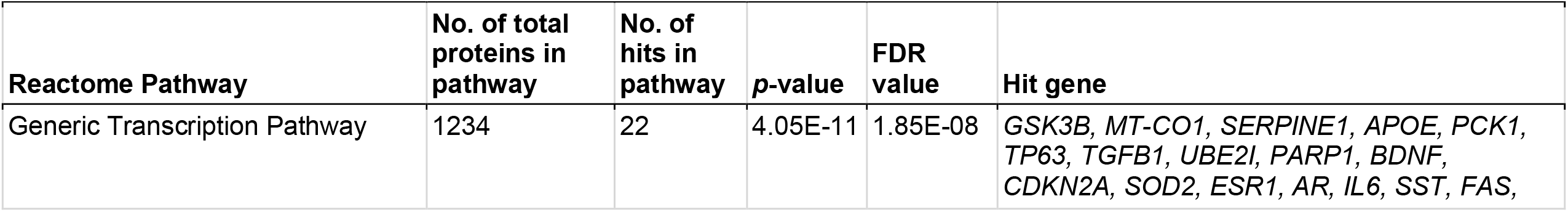
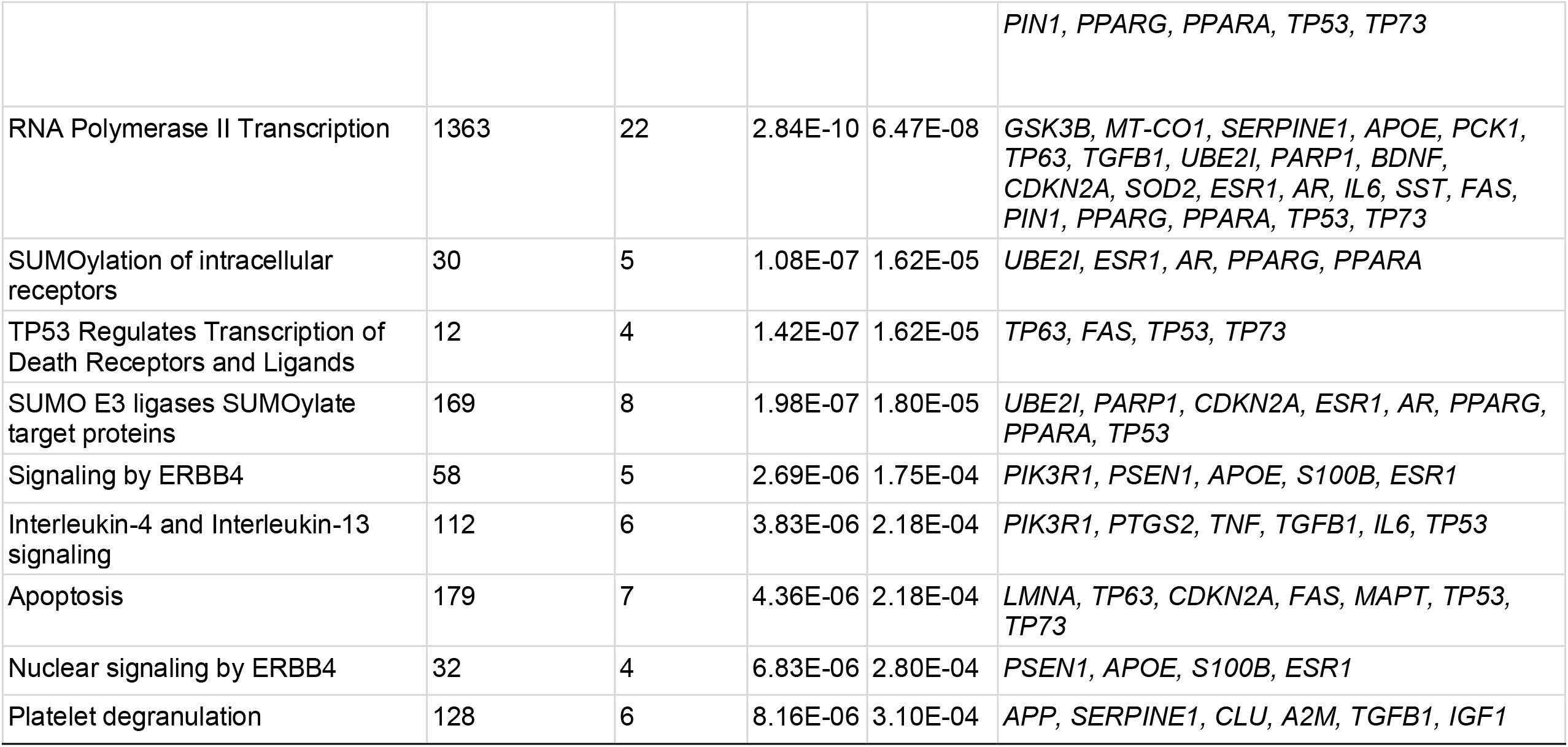
Reactome pathway enrichment analysis results - AD-AR Overlap genes.

The top result for the enriched pathway analysis was “Generic Transcription Pathway” (FDR = 1.85E-08; p = 4.05E-11), which was a sub-pathway topic under “RNA polymerase II transcription” (FDR 1.85E-08; p = 2.84E-10), which was a sub-pathway topic under the “Gene Expression” general pathway. Within “Generic Transcription Pathway”, the analysis highlighted “nuclear receptor transcription pathway” (FDR = 9.705E-04; p = 4.85E-05) and “TP53 regulates transcription of cell death genes” (FDR = 5.90E-04; p = 2.36E-05). “Generic transcription pathway” involves 1,234 proteins and 22 of those proteins were found to be shared in AD-AR overlap genes (FDR = 1.85E-08; p = 4.05E-11).

Pathways within the AD-AR overlap genes (Figures 2 and 3) for the “generic transcription pathway” included the following sub-pathways: “nuclear receptor transcription pathway” and “transcriptional regulation by TP53”. Under “SUMOylation” hit genes included the following sub-pathways: “SUMO E3 ligases SUMOylate target proteins”, “SUMOylation of intracellular receptors”, “SUMOylation of transcription factors”, and “SUMOylation of DNA damage response and repair proteins”.

**Figure 2.**
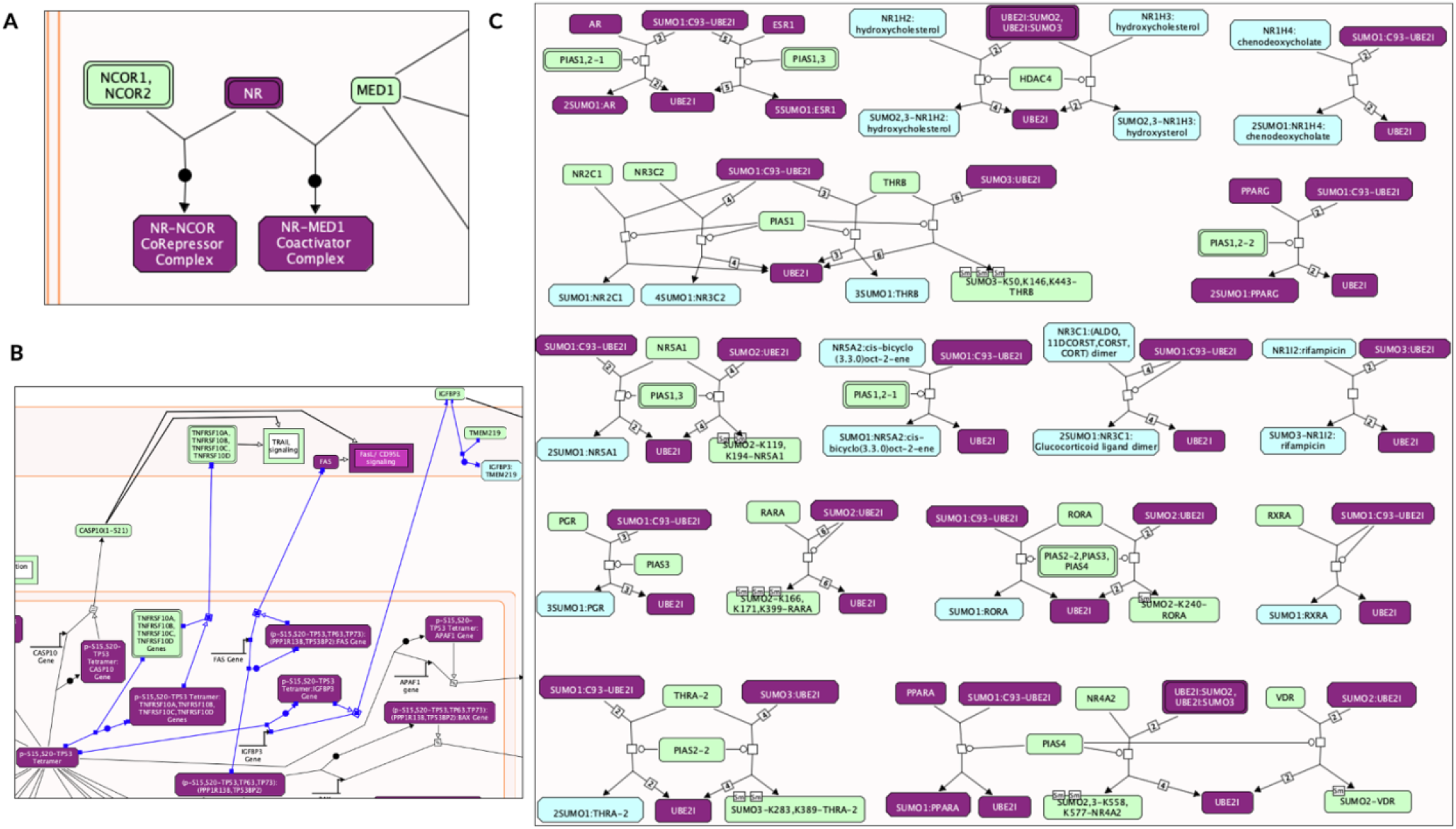
Enriched pathway analysis. The entities colored purple are hits within the positive gene list. Proteins are rectangles whereas elongated hexagons are complexes. (A) Generic transcription pathway. (B) TP53 Regulates the transcription of death receptors and ligands. (C) SUMOylation of intracellular receptors.

**Figure 3.**
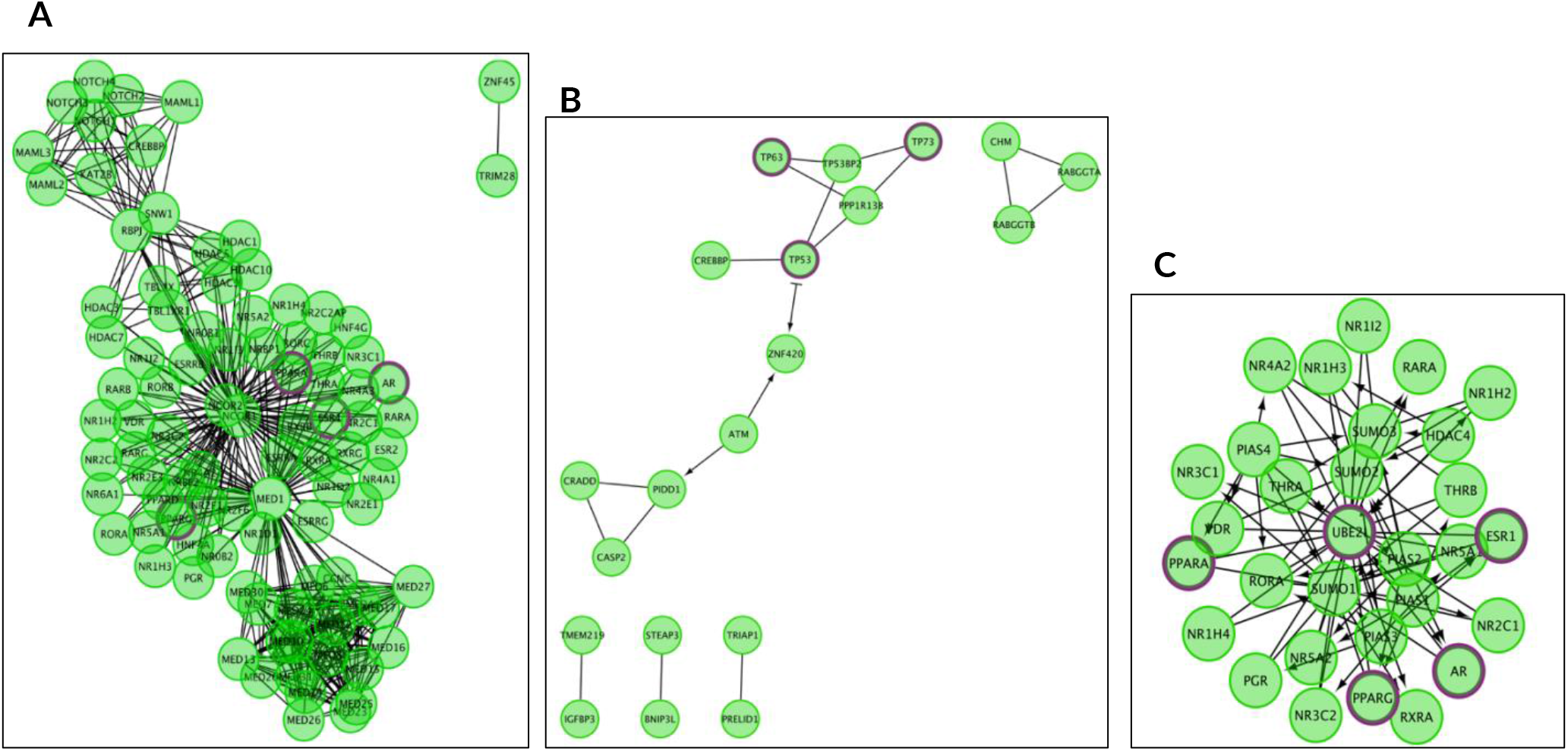
The Reactome pathways from figure 2 converted into a functional interaction network. Sub-pathways within the original pathway diagrams were extracted into the FI network as well. Hit genes are displayed in a thick purple border in the FI network view for a hit pathway. (A) Generic transcription pathway. (B) TP53 Regulates the transcription of death receptors and ligands. (C) SUMOylation of intracellular receptors.

### AD-Longevity overlap genes

43 genes of Longevity genes (12% of the genes) overlapped with AD genes (Table 6). The Reactome analysis of AD-longevity overlap genes identified 34 pathways, of which 12 pathways were validated within the threshold of p < 1.0E-05. The 12 pathways were in the following 4 Reactome general pathways: Immune System, Plasma lipoprotein assembly, remodeling, and clearance, Metabolism of vitamins and cofactors, and Signal Transduction. The top 10 results are shown in Table 7.

**TABLE 6.** Comparison of Longevity and AD Genes.

| TABLE 6 \| Comparison of Longevity and AD Genes |  |  |  |
| --- | --- | --- | --- |
| Longevity Genes | Positive AlzGene (AD) Genes | Overlap Genes | Overlapped in Longevity genes % |
| 357 | 356 | 43 | 12% |

**Table 7.** Reactome pathway enrichment analysis results - Longevity and AD Overlap genes.

| **Reactome Pathway** | **No. of total proteins in pathway** | **No. of Hits in Pathway** | **p-value** | **FDR value** | **Hit gene** |
| --- | --- | --- | --- | --- | --- |
| Interleukin-4 and Interleukin-13 signaling | 112 | 7 | 7.64E-08 | 2.32E-05 | *TNF, HMOX1, IL10, TGFB1, IL18, IL6, TP53* |
| Plasma lipoprotein assembly | 19 | 4 | 5.15E-07 | 7.82E-05 | *APOE, APOA1, APOA4, APOC1* |
| Plasma lipoprotein assembly, remodeling, and clearance | 71 | 5 | 3.69E-06 | 2.30E-04 | *CETP, APOE, APOA1, APOA4, APOC1* |
| Plasma lipoprotein remodeling | 32 | 4 | 4.02E-06 | 2.30E-04 | *CETP, APOE, APOA1, APOA4* |
| Chylomicron remodeling | 10 | 3 | 5.35E-06 | 2.30E-04 | *APOE, APOA1, APOA4* |
| Chylomicron assembly | 10 | 3 | 5.35E-06 | 2.30E-04 | *APOE, APOA1, APOA4* |
| HDL remodeling | 10 | 3 | 5.35E-06 | 2.30E-04 | *CETP, APOE, APOA1* |
| Signaling by Interleukins | 452 | 9 | 1.20E-05 | 4.57E-04 | *AGER, TNF, HMOX1, IL10, TGFB1, IL18, SOD2, IL6, TP53* |
| Retinoid metabolism and transport | 44 | 4 | 1.40E-05 | 4.61E-04 | *TTR, APOE, APOA1, APOA4* |
| Interleukin-10 signaling | 47 | 4 | 1.80E-05 | 5.29E-04 | *TNF, IL10, IL18, IL6* |

The top result from the enriched pathway analysis is “Interleukin-4 and Interleukin-13 signaling” (FDR = 2.32E-05; p = 7.64E-08) which is a sub-pathway of “signaling by Interleukins” (FDR 4.57E-04; p = 1.20E-05) and part of the “Immune System” general pathway. Out of the 112 proteins found to be involved in the “Interleukin-4 and Interleukin-13 signaling” pathways, 7 were shared by AD-Longevity overlap genes. Key hit genes involved with these pathways include TNF, IL10, IL18, and IL6. These genes are also shown to be involved in Interleukin 10 signaling. Interleukin pathways are shown in figures 4A and 5A.

**Figure 4.**
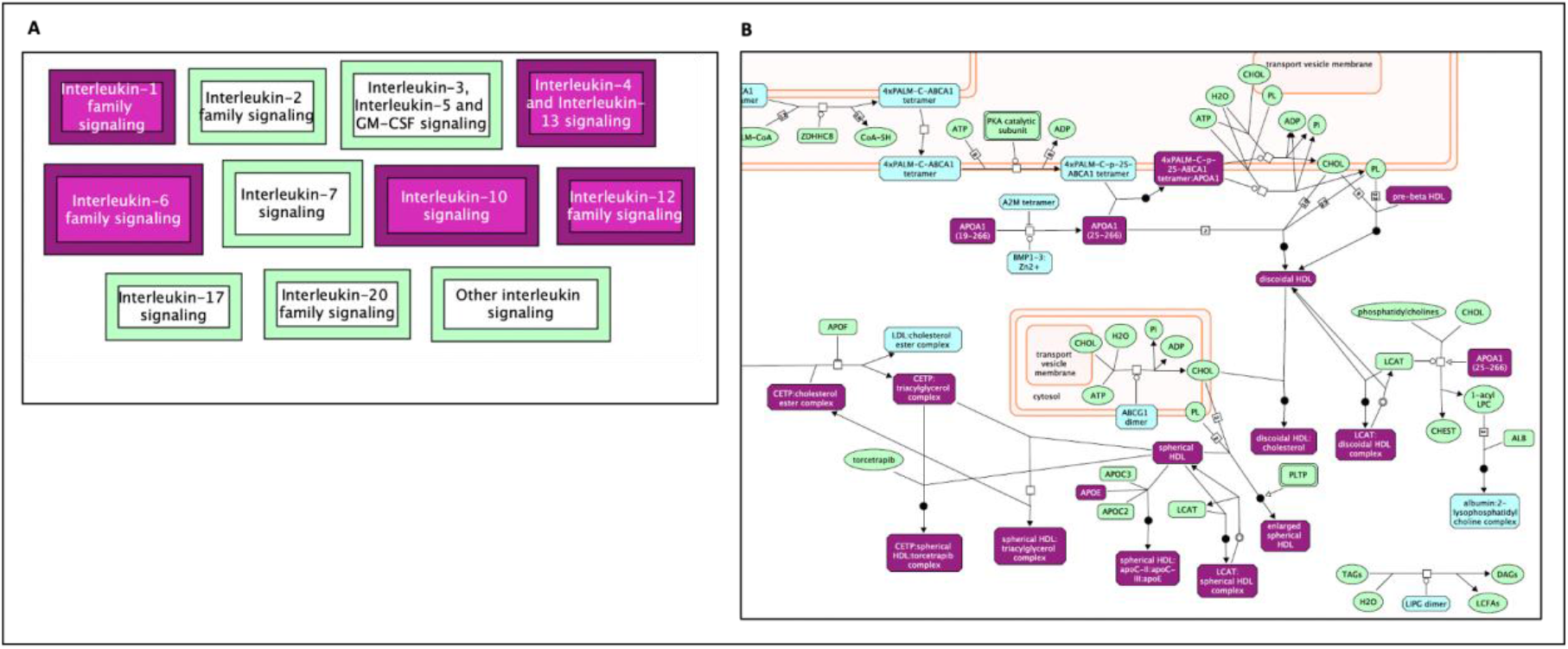
Enriched pathway analysis. The entities colored purple are hits within the positive gene list. Proteins are rectangles whereas elongated hexagons are complexes. (A) Generic transcription pathway. (B) TP53 Regulates the transcription of death receptors and ligands. (C) SUMOylation of intracellular receptors.

**Figure 5.**
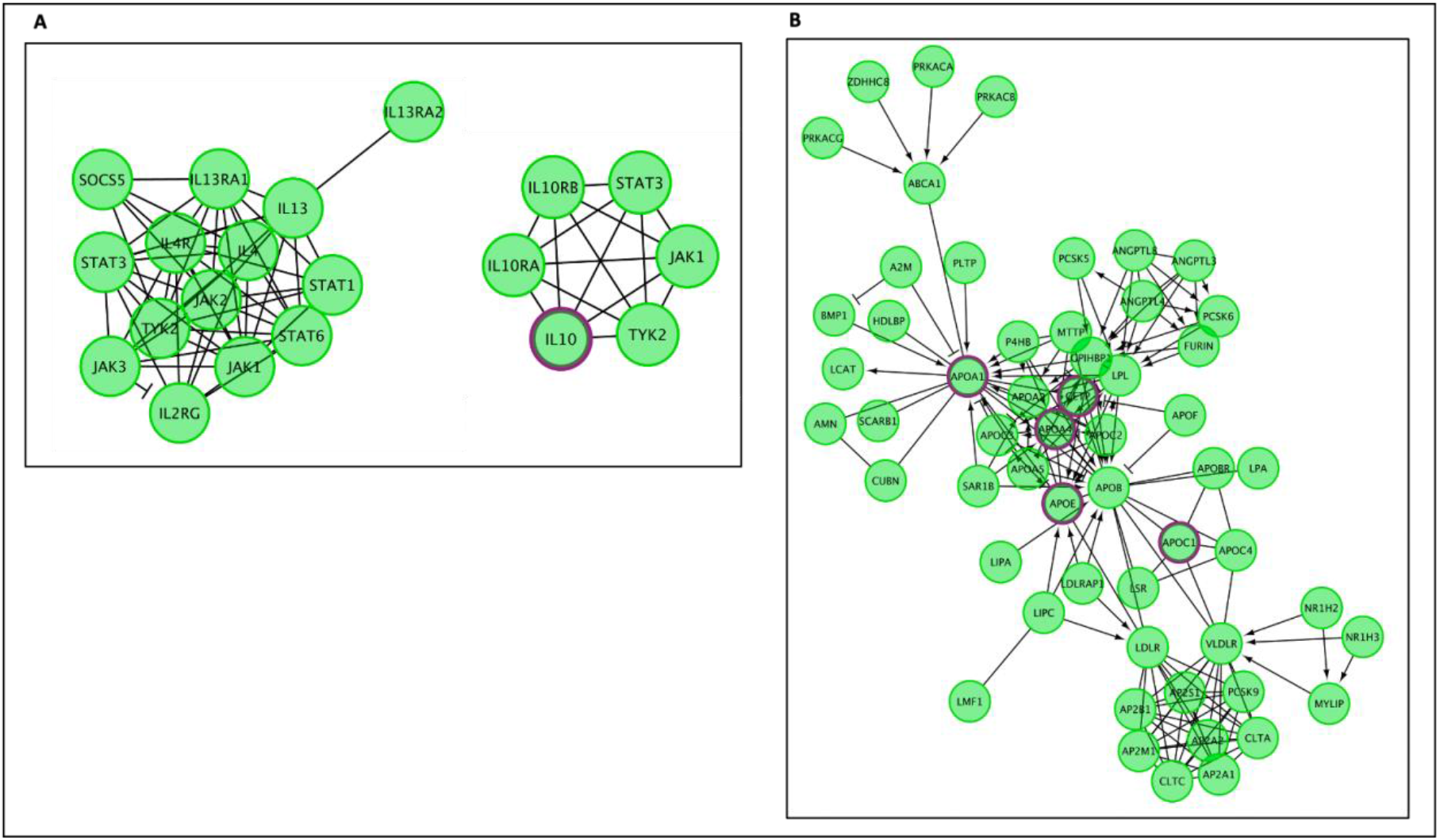
The Reactome pathways from figure 4 converted into a functional interaction network. Hit genes are displayed with a purple border. (A) FI Network for Diagram of Interleukin-4, Interleukin-13, and Interleukin-10 signaling, (B) FI Network for Diagram of Plasma lipoprotein assembly, remodeling, and clearance.

The second major Reactome group is “Plasma lipoprotein assembly, remodeling, and clearance” (FDR = 2.30E-04; p = 3.69E-06). Genes in this pathway (Figure 4b and 5b) incorporate the following significant sub-pathways: “plasma lipoprotein assembly” (FDR = 7.82E-05, p = 5.15E-07) which is a sub-pathway of “plasma lipoprotein remodeling”, “chylomicron remodeling”, “chylomicron assembly”, and “HDL remodeling.” The hit genes CEPT and APOE are present in all of these subcategories, highlighting their importance in this AD and longevity overlap dataset. Other top results included “Retinoid metabolism and transport”, which is a subset of “Metabolism of fat-soluble vitamins” and “NR1H2 and NR1H3-mediated signaling”, a subset of the “signal transduction” pathway. APOE is also a hit gene in these pathways.

## Discussion

This study identified a diverse range of biochemical pathways, using the Reactome analysis of each individual set of AD, AR, and Longevity genes. We compiled and removed redundancies of the pathways into comparable groups by highlighting the hallmarks of each subset and comparing each individual gene set to the pathways involved in the overlapping (AD-AR and AD-Longevity) gene sets. Figure 6 summarizes pathways involved in each subset, which are further discussed in the subsequent sections.

**Figure 6.**
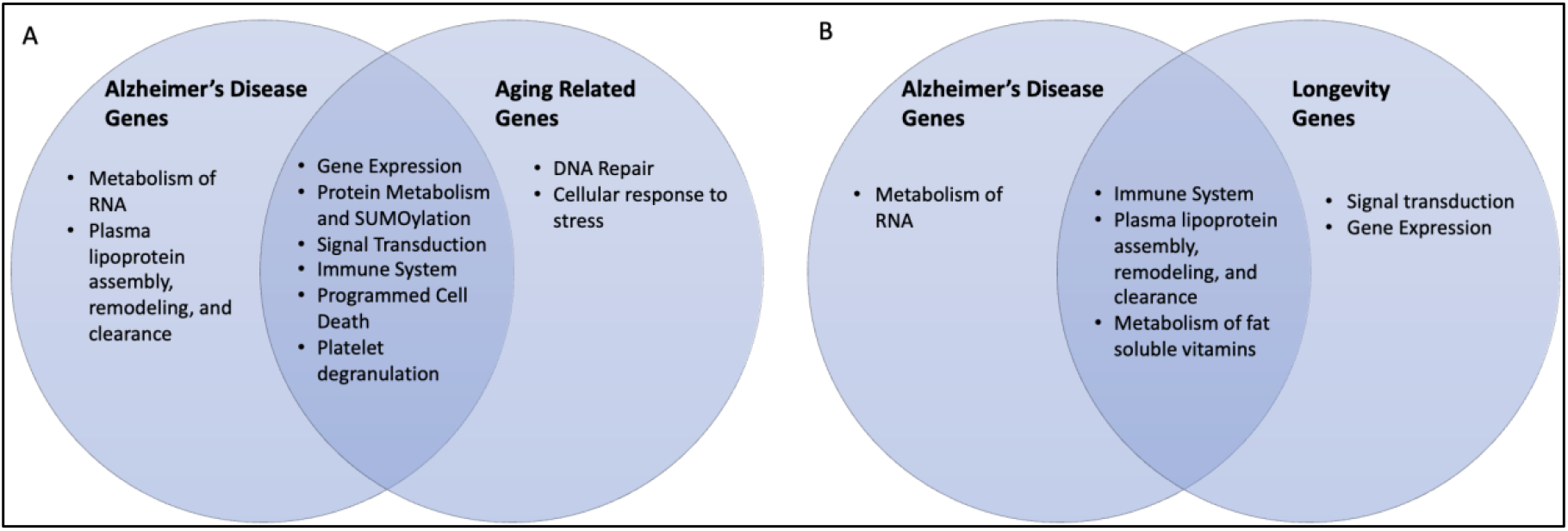
Venn diagrams of Overlapping Pathways: (A) Alzheimer’s genes and Aging Related genes and (B) Overlapping Alzheimer’s genes and Longevity genes.

### AD genes

Key genes involved in the AD subset are components of mitochondrial processing of RNA, lipoprotein metabolism, and interleukin signaling (Figure 7). This aligns with the mitochondrial cascade which proposes that mitochondrial function and change rates influence AD chronology. Reactome analysis has made mitochondrial genes the top hit when associated with AD genes alone, signaling its importance and the need for more data on possible interventions. Lipoprotein metabolism and interleukin signaling will be discussed in subsequent sections.

**Figure 7.**
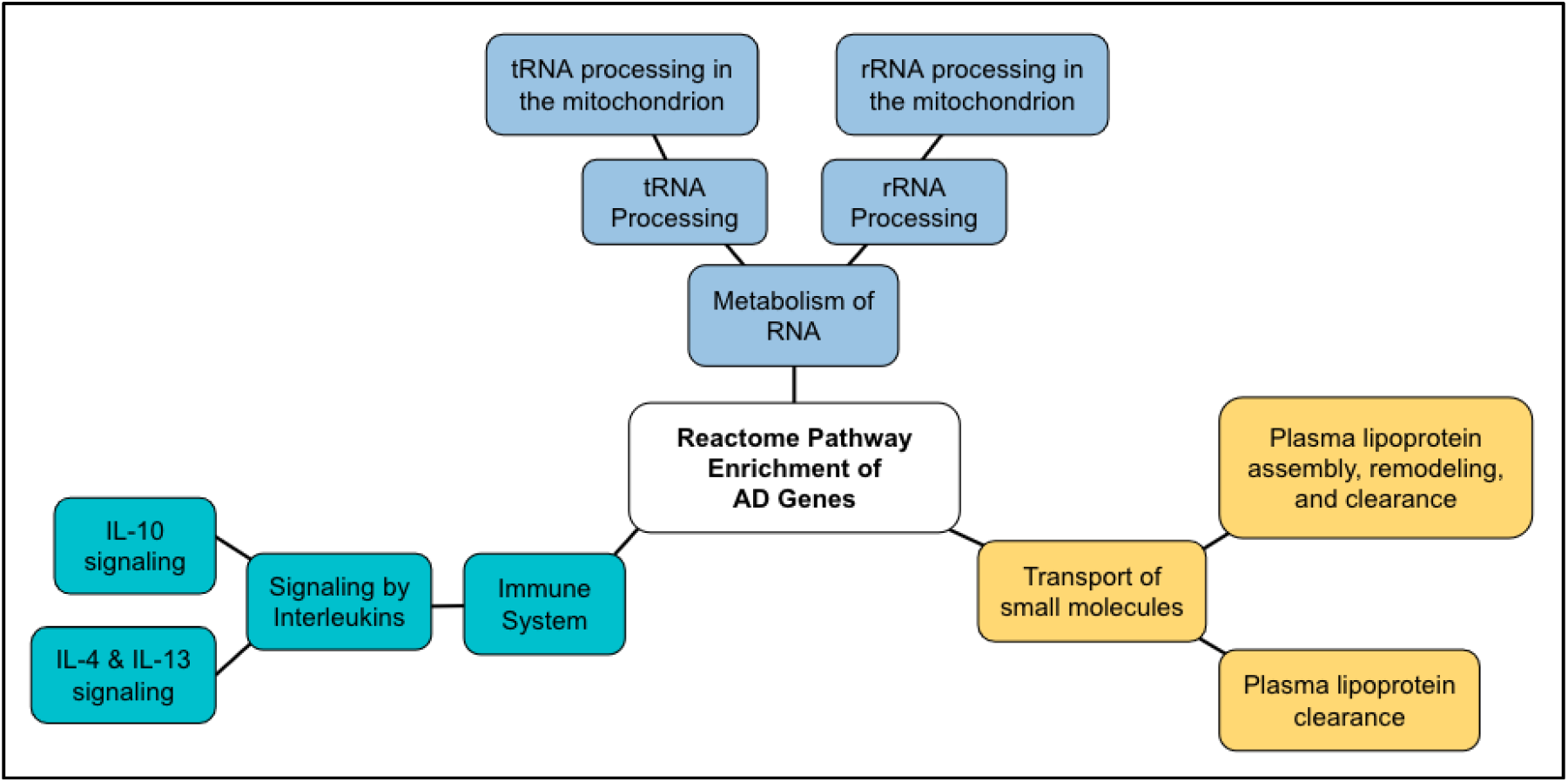
Diagram of Reactome Pathway Enrichment of AD Genes.

### AR genes

Key genes that are unique to the AR subset are components of DNA double-strand break (DSB) repair and cellular senescence (Figure 8). DSB can precipitate genomic rearrangements affecting multiple genes thus leading to much broader consequences when compared to other types of DNA mutations. This suggests that as humans age, DSB as well as other DNA repair mechanisms become less efficient and more-error prone (Gorbunova V et. al, 2016).

**Figure 8.**
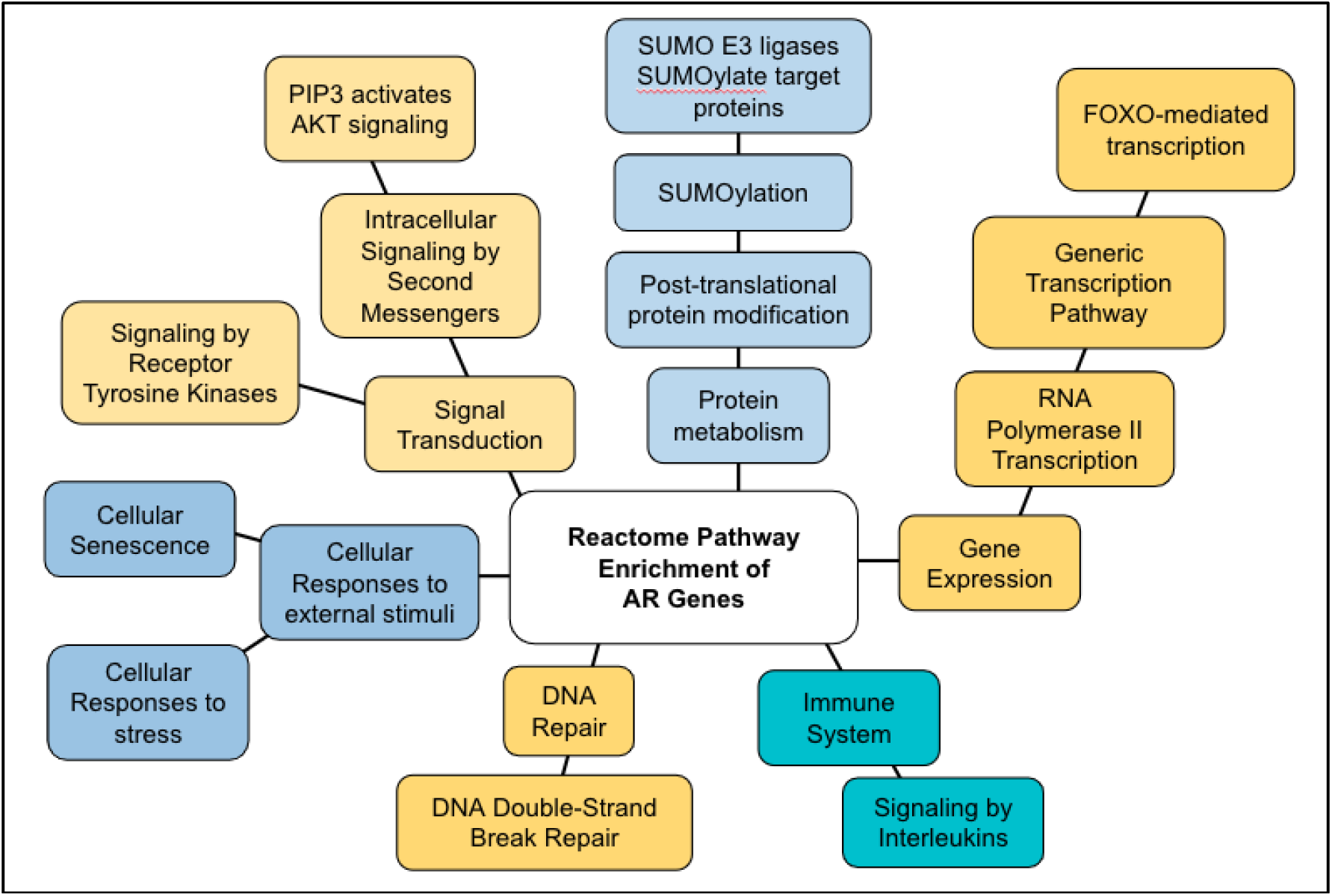
Diagram of Reactome Pathway Enrichment of AR Genes.

Cellular Senescence is a cellular response characterized by a stable growth arrest and other phenotypic alterations that play a role in normal development, tissue homeostasis, and preventing tumor progression. It has been implicated as a major cause of the age-related disease (McHugh et. al, 2018). This suggests that further study of cellular senescence can lead to novel therapies for age-related diseases.

### Longevity genes

Key genes involved in longevity are components of the PIP3-ATK and mTOR pathways (Figure 9). PIP3-AKT is a part of the insulin and IGF-1 signaling pathway which regulates FOXO (forkhead box O), a subset of a large family of transcription factors which have been shown to be involved in many cellular processes such as cell growth and survival, especially FOXO3 and FOXO1 genes (Martins et al., 2015). The mechanistic target of rapamycin (mTOR), which is also a downstream target of the insulin/PIP3/AKT pathway, is a protein kinase in a highly conserved pathway that senses nutrients and other environmental signals and coordinates several fundamental cellular responses, such as cell growth and proliferation, and has been linked to the physiological process of aging. (Saxton and Sabatini, 2017). Previous studies have shown that inhibition of mTOR enhances longevity and decreases aging and age-related disease in model organisms (Weichhart 2018). The transcription factor TP53 which encodes tumor suppressor p53 is involved in several aging-related pathways such as apoptosis and senescence and has also been shown to influence insulin/mTOR signaling which can contribute to longevity (Feng et al., 2011).

**Figure 9.**
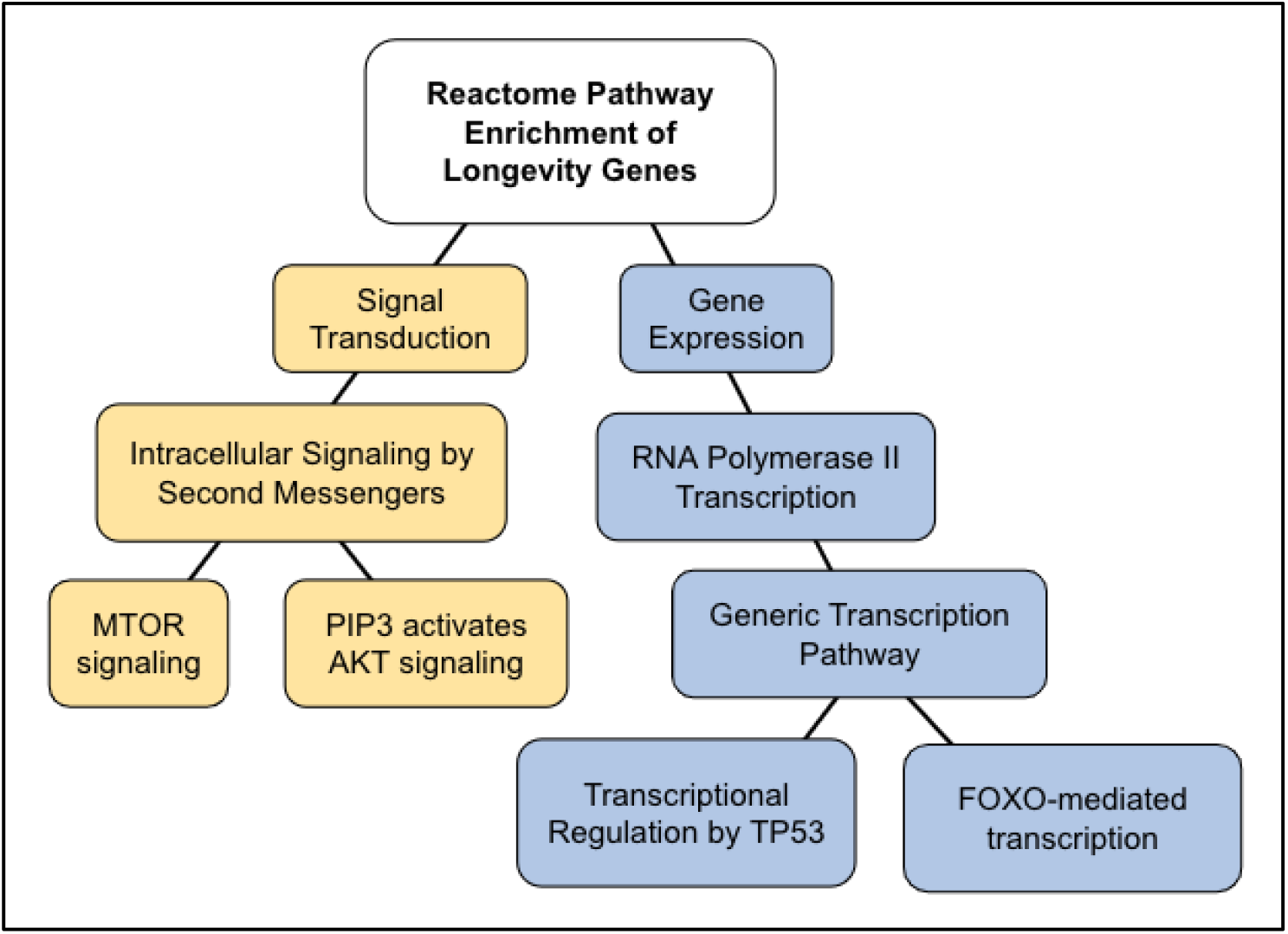
Diagram of Reactome Pathway Enrichment of Longevity Genes.

### Comparing AR Pathways with Longevity Pathways

Based on our Reactome analysis, there are shared biological pathways among AR and Longevity genes, including the immune system and cytokine signaling, signal transduction, and gene expression are involved in the balance between aging and longevity. Aging also has components of impaired DNA repair, metabolism of proteins, as well as cellular senescence. We have compared the pathways involved in AR and Longevity genes, which represent the balance of shared pathways (Figure 10).

**Figure 10.**
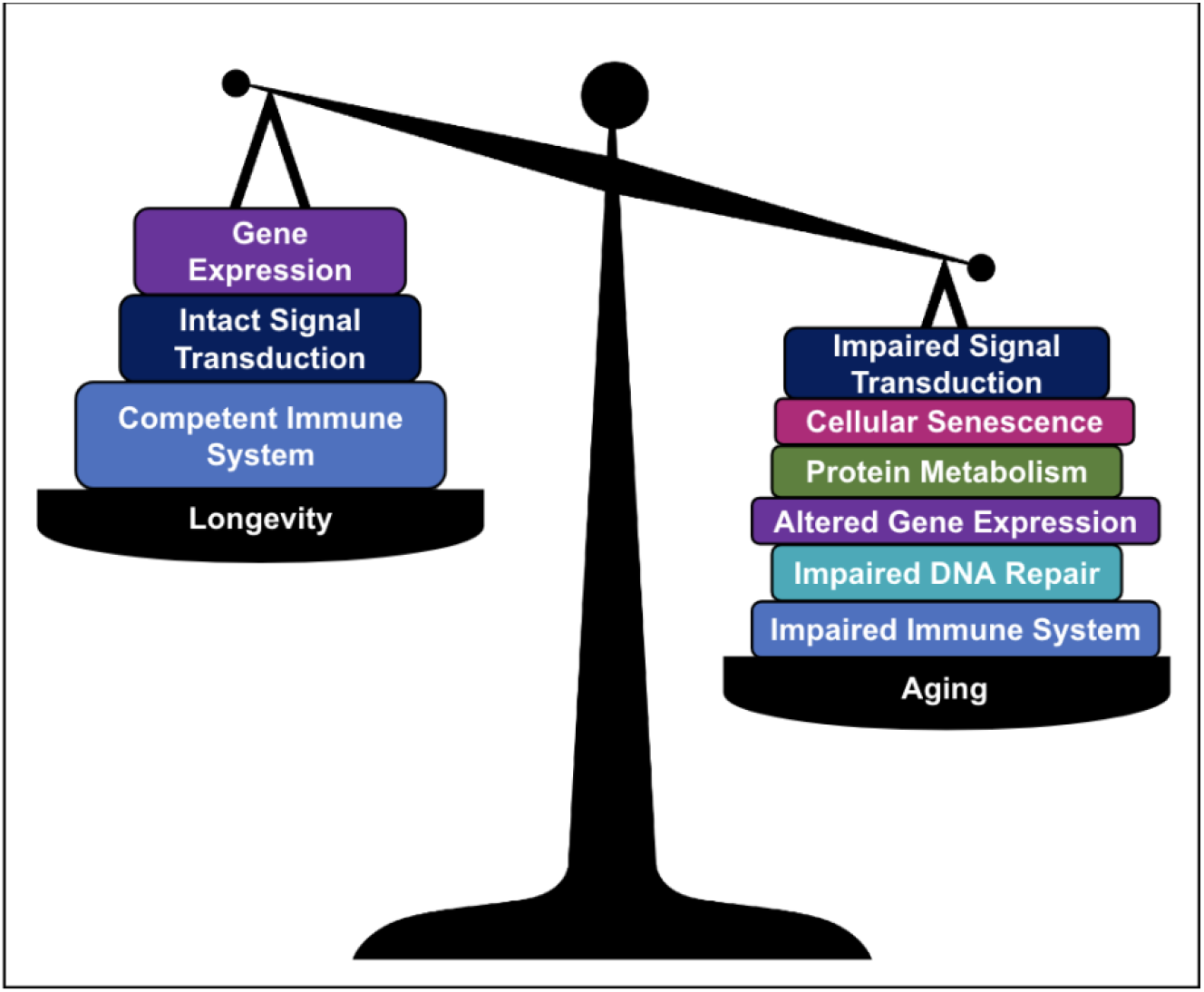
A schematic illustrating proposed genetic pathways that contribute to aging and longevity.

### AD-AR overlap genes

Although the gene ontology for the AD-AR Overlap genes demonstrated involvement in a broad range of biological pathways, concurrent with previous studies, some of our most significant associations involve TP53, FOXO, and SUMOylation (Table 4 and Figure 11). The TP53 gene provides instructions for making a prominent tumor suppressor protein called tumor protein p53 which regulates DNA repair and cell division. This adds to the consistent support that p53 represents a potential peripheral biomarker that could detect AD at its earliest stages (Abate et al., 2020).

**Figure 11.**
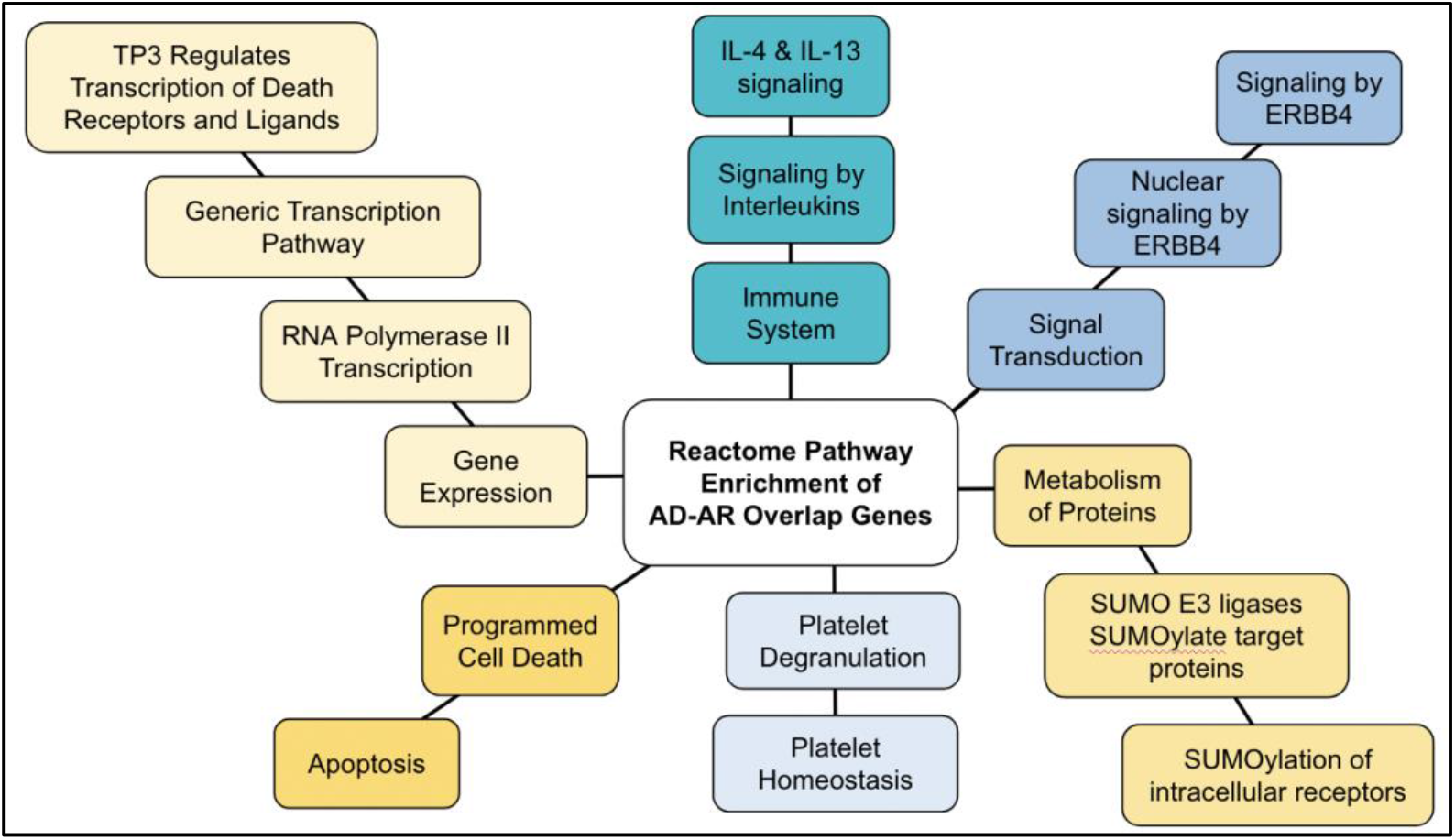
Diagram of Reactome Pathway Enrichment of AD-AR Overlap Genes

FOXO proteins are a subgroup of the Forkhead family of transcription factors involved in the regulation of metabolism, regulation of reactive species, and regulation of cell cycle arrest and apoptosis. This adds to the support that there has been increased plaque load and core plaque size, commonly found in AD progression, in the cortex of mice with FOXO3 deficiency (Shuqi et al., 2019).

Histopathologically, AD features insoluble aggregates of two proteins in the brain, amyloid-β (Aβ) and tau aggregates - both of which have been linked to the small ubiquitin-like modifier (SUMO). This adds to the support that early studies have indicated that the SUMO system is likely altered with AD-type pathology (Lee et al., 2013).

### AD-Longevity overlap genes

Significant pathways generated from the AD-Longevity overlap set include cytokine signaling in the immune system and lipoprotein metabolism, which is related to the metabolism of fat-soluble vitamins and NR1H2 and NR1H3-mediated signaling. (Figure 12).

**Figure 12.**
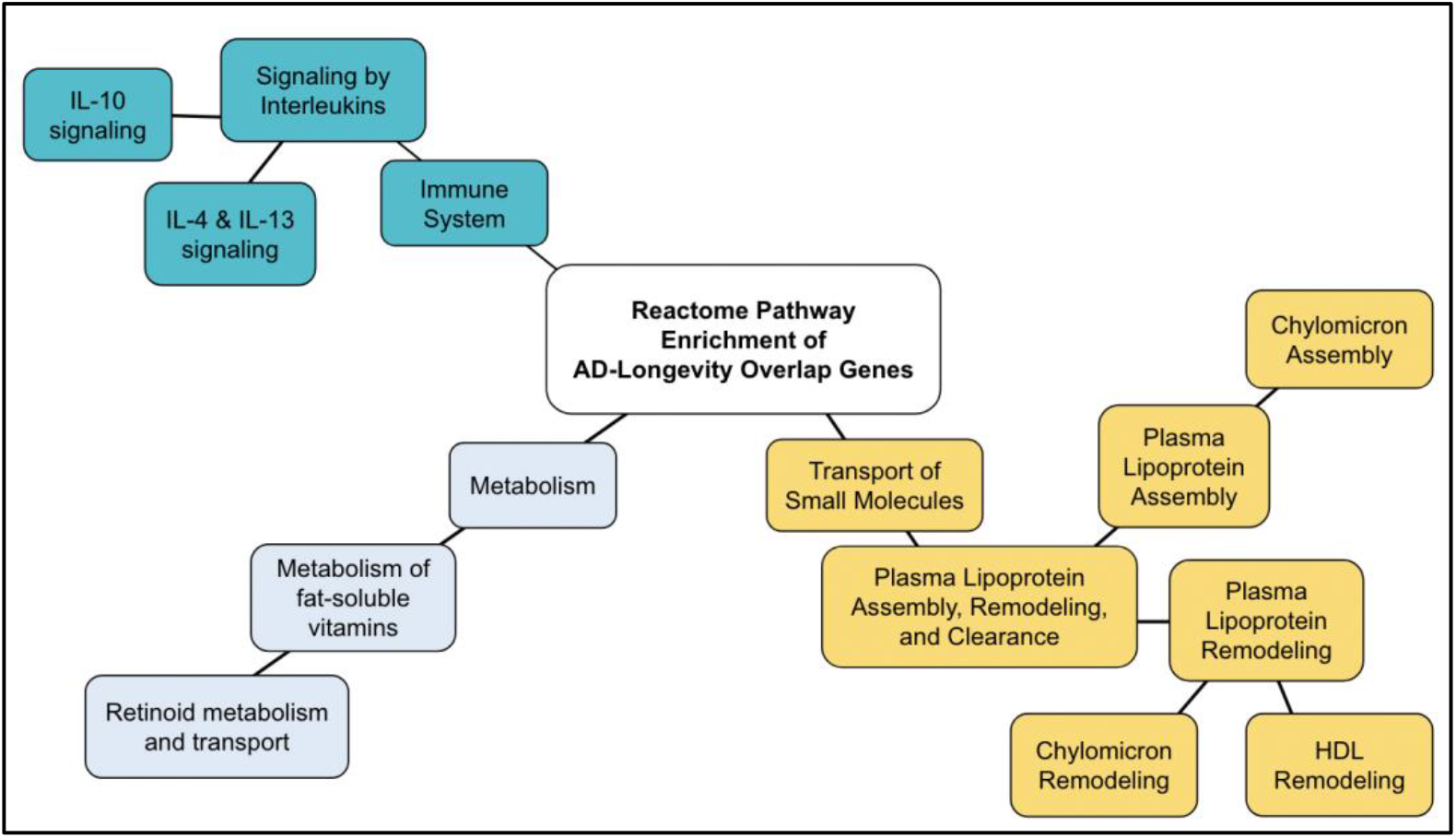
Diagram of Reactome Pathway Enrichment of AD-Longevity Overlap Genes.

Cytokines are known to play an integral role in regulating inflammatory pathways, including neuroinflammation in Alzheimer’s disease. Aß plaques have been shown to increase levels of proinflammatory cytokines IL6 and TNF alpha-among others, leading to a vicious cycle of cytokine-derived inflammation and plaque accumulation (Rubio-Perez 2012). Proinflammatory cytokines TNF alpha and especially IL6 have been shown to become more prevalent with age. Thus genetic differences in these cytokines could serve as a potential connection to either longevity or aging and inflammatory age-related diseases (Singh 2010). It has been theorized that in longer-lived individuals, the polymorphisms that express lower IL6 levels would be negatively correlated, but studies have had mixed results (Guiliani et al., 2018) (Di Bona D., 2009), (Wei GZ, 2016). IL4 is anti-inflammatory in the brain and has been shown to potentially counter the inflammatory processes in age-related diseases in mice, although detailed mechanisms are not yet known (Gadani 2012). Our present study reveals the importance of the interplay and balance of cytokines, especially IL4 and IL6, which would be a potential area for further research.

The second category in the AD-longevity overlap genes is plasma lipoprotein metabolism. Interestingly, lipoprotein metabolism was not a top result when analyzing the longevity genes alone, but is significant in the overlap group. This reiterates the importance of lipoprotein metabolism specifically in the context of its relation to AD. Two genes are important in this pathway: Apolipoprotein E (APOE) and cholesteryl ester transfer protein (CEPT).

Polymorphisms of APOE are an established risk factor for developing AD by enhancing Aß-led inflammation and deterioration (Yamazaki 2019). Polymorphisms of CEPT are potential candidates for risk factors for developing AD but results have been discordant. (Arias-Vásquez et al., 2007). Despite being shown to increase the risk of developing AD, these genes are difficult to target or change, since they are needed for normal lipid homeostasis. Further, variants could be evolutionarily beneficial earlier in life, an example of antagonistic pleiotropy (Tuminello and Han, 2011). Thus, future studies should emphasize the interplay between genes and lifestyle throughout a person’s lifetime.

### Technical advantages and limitations

Gene set enrichment analysis (GSEA) is a computational method that is useful to interpret the biological functions of a gene set with statistical confidence. Reactome incorporates GSEA embedded with the knowledge database that covers more than 100 bioinformatics resources. The Reactome outputs include statistical confidence levels with p-values and false discovery rate, which raises the confidence level of evidence. The gene enrichment algorithms use a score-ranked list and compare it with random rankings (Subramanian et al. 2005), which provides a reasonable level of statistics with a p-value and false discovery rate. Thus, we suggest that the results from this study represent the most current genetic hallmarks involved in AD, aging, and longevity. In fact, this study provides a comprehensive update of the genomic map of the AR genes reported nearly two decades ago (de Magalhaes and Toussaint, 2004).

However, the results from GSEA should be viewed as living knowledge, which requires updates until the knowledge is saturated and complete. A potential limitation is that connections among genes and annotations may be influenced by previous studies, which can be minimized by peer review, systematic review, and meta-analysis. Although poor study design and controls in a GWAS study may influence overall annotations, it is likely that GSEA and the review systems minimize the impact of such a study. We also used a stringent condition (p < 1.00E-05), which is backed up by the false discovery rate < 1.00E-03. Additionally, knowledge databases, such as Reactome (https://reactome.org/) and KEGG (https://www.genome.jp/kegg/), process the data rigorously with continued updates, which should reduce incorporating experimental errors.

Another limitation is manually-curated annotation nomenclatures, some of which may not be easy to understand. For example, stress resistance to a variety of stresses (i.e., multiplex stress resistance) is a component of longevity (Murakami and Johnson, 1996; Murakami, 2006; Miller et al., 2017), which cannot be identified using the current annotation and thus remains to be investigated.

### Future Direction

In this present study, we investigated and interpreted the gene profiles of aging and longevity and their association with Alzheimer’s disease genes. We used the Reactome analysis which provides gene set enrichment analysis to provide confidence levels for each genetic pathway. Our results suggest overlapping pathways that involve TP53, FOXO, protein metabolism (SUMO), cytokine balance, and lipoprotein metabolism. The genetic hallmarks identified in this study provide unexpectedly broad mechanisms, suggesting a wide variety of implications in the field of aging. Manually-curated annotations may need to be optimized to understand AD, aging, and longevity more accurately. Further studies about environmental and lifestyle factors may strengthen the knowledge about the development of Alzheimer’s disease and age-related comorbidities (Le et al., 2021; Li and Murakami, 2022). The studies may link aging, mid-life common diseases, and Alzheimer’s disease (Murakami et al., 2011; Murakami, 2013). The predictive contribution of each genetic variant remains modest for LOAD (Harold et al., 2009), we predict that the overviews of the gene sets, including those in this study, continue to be important not only for the understanding of AD but also for patients and education. Previously we have proposed to include feedback from patients in research when studying health (Murakami and Halperin, 2014) and provided an example (Murakami, 2016). We incorporated their feedback that there might be a more meaningful approach to AD patients than simply identifying the AD genes. The discussion prompted us to explore related genes and extracted the biological hallmarks shared among aging, longevity, and AD. The genetic interaction networks among aging, longevity, and AD provide the extraction and translation of the gene information into the hallmarks as well as are the key to developing effective treatments for AD. We hope to stimulate basic science research open to patients, the community, and education.

## Data Availability

All data produced in the present work are contained in the manuscript

## Acknowledgment

This work was initiated at the Program of the Master of Science in Medical Health Sciences, Touro University California. We thank all the members of the Murakami laboratory at Touro University-California for the helpful discussion.

## Notes

### Competing Interest Statement

The authors have declared no competing interest.

### Funding Statement

This study did not receive any funding

### Author Declarations

The human gene data used were retrieved from: GenAge database (www.genomics.senescence.info) LongevityMap database (www.genomics.senescence.info/longevity/) and PubMed and NCBI database (https://www.ncbi.nlm.nih.gov/)

